# Regional spread of bla_NDM-1_-containing *Klebsiella pneumoniae* Sequence Type 147 in post-acute care facilities

**DOI:** 10.1101/2021.03.16.21253722

**Authors:** Zena Lapp, Ryan Crawford, Arianna Miles-Jay, Ali Pirani, William E. Trick, Robert A. Weinstein, Mary K. Hayden, Evan S. Snitkin, Michael Y. Lin

## Abstract

**Background:** Carbapenem-resistant *Enterobacterales* (CRE) harboring bla_KPC_ have been endemic in Chicago-area healthcare networks for more than a decade. During 2016-2019, a series of regional point prevalence surveys identified increasing prevalence of bla_NDM_-containing CRE in multiple long-term acute care hospitals (LTACHs) and ventilator-capable skilled nursing facilities (vSNFs). We performed a genomic epidemiology investigation of bla_NDM_-producing CRE to understand their regional emergence and spread.

**Methods:** We performed whole-genome sequencing on NDM+ CRE isolates from four point-prevalence surveys across 35 facilities (LTACHs, vSNFs, and acute care hospital medical intensive care units) in the Chicago area and investigated the genomic relatedness and transmission dynamics of these isolates over time.

**Results:** Genomic analyses revealed that the rise of NDM+ CRE was due to the clonal dissemination of an ST147 *Klebsiella pneumoniae* strain harboring bla_NDM-1_ on an IncF plasmid. Dated phylogenetic reconstructions indicated that ST147 was introduced into the region around 2013 and likely acquired NDM around 2015. Analyzing genomic data in the context of patient transfer networks supported initial increases in prevalence due to intra-facility transmission in certain vSNFs, with evidence of subsequent inter-facility spread to connected LTACHs and vSNFs via patient transfer.

**Conclusions:** We identified a regional outbreak of bla_NDM-1_ ST147 that began in and disseminated across Chicago area post-acute care facilities. Our findings highlight the importance of performing genomic surveillance at post-acute care facilities to identify emerging threats.

## Introduction

Carbapenem-resistant *Enterobacterales* (CRE) represent an urgent antibiotic resistance threat due to their resistance to first-line antibiotics and transmissibility in healthcare settings [1,2]. The emergence of epidemic lineages of CRE that are resistant to nearly all antibiotics and that cause infections with high mortality rates, such as *Klebsiella pneumoniae* carbapenemase (KPC) containing *Klebsiella pneumoniae* (KPC-Kp) sequence type (ST) 258 [3], has further escalated the need for more effective strategies to interrupt CRE transmission. Most interventions to prevent the spread of CRE and other healthcare-associated antibiotic resistance threats have been implemented at the level of individual healthcare facilities. However, there is now a multitude of evidence that the frequent movement of colonized and infected patients among regional healthcare facilities necessitates regional surveillance and infection prevention strategies [4].

Long-term acute care hospitals (LTACHs) and ventilator-capable skilled nursing facilities (vSNFs) are potentially high-impact settings for implementation of regional CRE surveillance and infection prevention interventions [5,6]. Patients in these facilities have been shown to be colonized with CRE at high rates, likely due to a combination of their chronic severe illness, long lengths of stay, and high rates of prior or on-going antibiotic exposure. Modeling and epidemiologic studies have suggested that the high CRE prevalence in LTACHs in particular has a significant impact on connected healthcare facilities with which they share patients [7,8]. Currently, less is known about the regional influence of vSNFs, although the even longer lengths of patient stay and more limited resources for infection prevention indicate that they might also be important settings in regional amplification of antibiotic resistance.

A bundled infection prevention intervention [9] (Chicago PROTECT [10]) was initiated in July 2017 to control CRE in Chicago-area healthcare facilities, including in vSNFs and LTACHs. Serial point prevalence surveys conducted to monitor the impact of the intervention demonstrated that KPC-Kp levels remained stable across regional facilities. However, during the intervention period, New Delhi metallo-beta-lactamase (NDM) containing CRE prevalence unexpectedly increased in a subset of surveyed vSNFs. Here, we applied whole-genome sequencing to understand the molecular and epidemiological basis for the increase in NDM prevalence in the region.

## Materials & Methods

### Study isolates and metadata

Starting from October 2016 until July 2019, 24 medical ICUs in 24 short term acute care hospitals, 6 LTACHs, and 8 vSNFs in the Chicago region were invited to participate in serial one day point prevalence surveys of residents on their skilled nursing and ventilator wards. Medical ICUs were surveyed once in 2016-2017; vSNFs and LTACHs were surveyed every 6-12 months. Local staff obtained a rectal swab sample from each patient and collected de-identified patient information assessed at time of survey (age up to 90 years of age, sex, respiratory support status, length of stay, contact precautions status, facility awareness of resident CRE status). Swabs were processed at a central lab within 6 hours of collection. Overnight growth from MacConkey agar plates was screened for 5 carbapenemase gene families (KPC, NDM, VIM, IMP & OXA-48) using multiplex PCR assays (Acuitas® MDRO gene test and Acuitas® Resistome test, OpGen, Gaithersburg, MD during 2016-2017; Xpert® Carba-R, Cepheid, Sunnyvale, CA during 2018-2019).

For all analyses, only the first isolate of a given ST (for *K. pneumoniae* isolates) or species (for all other species) was used for each patient. We used a Fisher’s exact test to test for the statistical significance of the difference in NDM or KPC prevalence between the first and last surveys. Fisher’s exact p-values were corrected using the Benjamini-Hochberg method.

### Whole-genome sequencing

Genomic DNA was extracted from cultures derived from single sub-cultured colonies. Genomic libraries were prepared with NEBNext Ultra DNA library prep kit and sequenced at the University of Michigan Advanced Genomics Core on an Illumina NovaSeq 6000. All sequenced isolates have been deposited under BioProject PRJNA686897.

### Genomic analysis

We processed whole-genome sequences [11,12] and identified in-silico multi-locus sequence types [13,14], generated and annotated assemblies [12,15–20], called single-nucleotide variants (SNVs) [21–28], identified phylogenetic clustering of facilities [8], and calculated pairwise SNV distances between isolates [29]. Reference-based whole-genome alignments of study and public ST147 isolates [30–40] were used to generate a phylogenetic tree using IQ-TREE v1.6.12 [41,42]. We inferred ancestral dates of the phylogeny with the R package BactDating v1.0.12 [43–45]. NDM-containing plasmids were identified from publicly available complete plasmids [29,29,46–48,21,49,50]. See supplemental methods for details of the genomic analysis.

### Determining patient flow between facilities

We constructed a patient transfer matrix of the Chicago metropolitan region using the Centers for Medicare and Medicaid Services’ minimum data set, Medicare Provider Analysis and Review [MEDPAR] limited data set, Medicaid Analytic eXtract Data from 2010-2012. Using the patient transfer matrix, we constructed a directed weighted patient transfer network of healthcare facilities in the Chicago area using R igraph v1.2.6 [51], including all healthcare facilities in the study, and patient flow was determined as in [8]. See supplemental methods for details about calculating patient flow.

### Data analysis and visualization

Data analysis and visualization was performed in R v4.0.2 [52]. Data visualization used the following packages: tidyverse v1.3.0 [53], pheatmap v1.0.12 [54], lubridate v1.7.9.2 [55], tidytree v0.3.3 [56], treeio v1.12.0 [57], ggtree v2.2.4 [58,59], ggplotify v0.0.5 [60], ggnewscale v0.4.4 [61], and cowplot v1.1.0 [62]. Code for analysis and visualization can be found here: https://github.com/Snitkin-Lab-Umich/ndm-st147-chicago-ms.

### Ethical Review

Bacterial isolates and de-identified clinical metadata were collected under a prior surveillance project that underwent ethical review at the CDC and was determined to be a nonresearch activity (public health surveillance). The project was also evaluated independently at each participating healthcare facility and either deemed a public health assessment or human subjects research and approved by local review boards where applicable.

## Results

### Prevalence of NDM, but not KPC, increased over time in certain vSNFs that are not closely connected by patient transfer

We first detected the presence of NDM+ CRE isolates in vSNFs and LTACHs during a regional point prevalence survey conducted in 2017 (survey 13; **Fig 1A**). To more closely monitor the potential emergence and spread of NDM in the region we performed three additional surveys for CRE colonization in regional vSNFs and LTACHs, each 6 months apart (surveys 13-1, 13-2, and 13-3; **Table 1**). We found that, while the prevalence of KPC+ CRE remained relatively stable over time in all facilities, the prevalence of NDM+ CRE increased in three vSNFs not closely connected by patient transfer (Fisher’s exact p < 0.05 for vSNFs J, K, and L; **Fig 1B, S1, S2; Table S1**).

**Figure 1:**
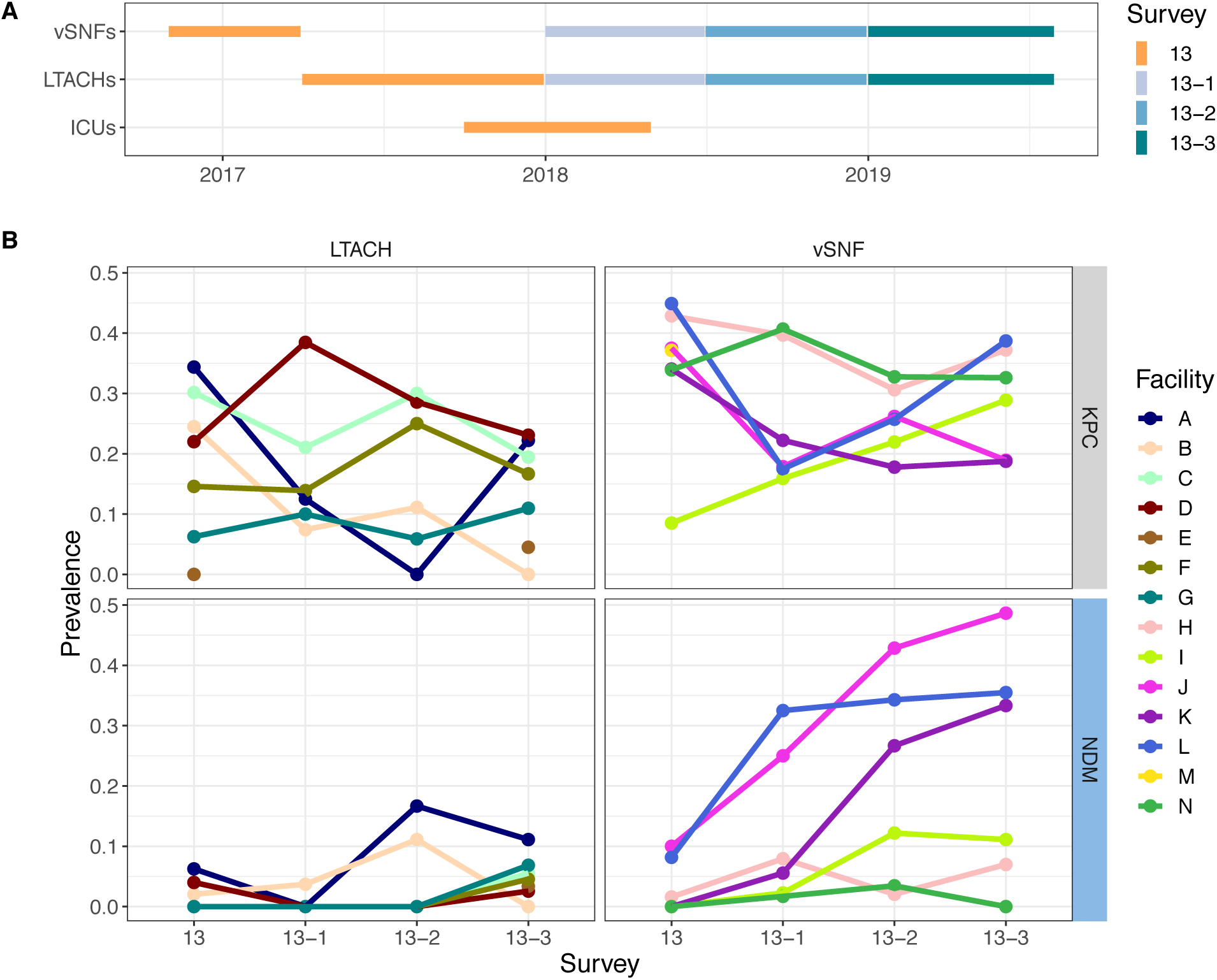
Prevalence of NDM is increasing over time in certain vSNFs. (A) Time window of when facilities were tested for each survey. (B) Proportion of NDM and KPC positive samples across surveys and facilities. vSNF O was not included as there was uneven sampling across surveys. vSNF = ventilator-capable skilled nursing facility. LTACH = long-term acute care hospital, ICU = intensive care unit. ICUs are not shown in panel B because of very low prevalence (see Table 1).

**Table 1:** Point prevalence survey results from ICUs, LTACHs, and vSNFs

|  | ICU | LTACH | vSNF |
| --- | --- | --- | --- |
| **Number of facilities** | 20 | 7 | 8 |
| **Age, mean yrs (SD)** | 62 (17) | 62 (15) | 60 (15) |
| **Male, n (%)** | 119 (56) | 481 (56) | 622 (55) |
| **Length of stay, median days (IQR)** | 5 (3-10) | 19 (12-34) | 127 (32-409) |
| **Mechanical ventilation, n (%)** | 102 (48) | 279 (22) | 472 (33) |
| **Tracheostomy collar, n (%)** | 0 (0) | 186 (22) | 378 (33) |
| **Contact precautions, n (%)** | 58 (27) | 533 (62) | 672 (59) |
| **Carbapenemase gene:** |  |  |  |
| **KPC, n (%)** | 11 (5) | 136 (16) | 357 (31) |
| **NDM, n (%)** | 2 (1) | 23 (3) | 145 (13) |
| **OXA-48, n (%)** | 1 (0) | 0 (0) | 1 (0) |
| **IMP, n (%)** | 0 (0) | 1 (0) | 0 (0) |
| **VIM, n (%)** | 0 (0) | 4 (0) | 66 (6) |
| **Any carbapenemase gene, n (%)** | 14 (7) | 152 (18) | 476 (42) |
| **Of carbapenemase-positive:** |  |  |  |
| **Carbapenemase-positive with contact precautions, n/N (%)** | 9/14 (64) | 127/152 (84) | 362/476 (76) |
| **Carbapenemase-positive known to facility, n/N (%)** | 6/14 (43) | 79/152 (52) | 301/476 (63) |

### The majority of NDM+ isolates are *K. pneumoniae* ST147 and carry bla_NDM-1_ on an IncF plasmid

To understand the molecular basis for the increase in NDM+ CRE we performed whole-genome sequencing on all CRE isolates from survey 13 and NDM+ isolates from the subsequent three follow-up surveys. We found that the presence of NDM was highly correlated with the presence of a suite of genes present on an NDM+ IncF plasmid isolated from *K. pneumoniae* [63] (**Fig S3**). Most isolates containing the IncF plasmid were bla_NDM-1_ *K. pneumoniae* ST147; however, one bla_NDM-1_ *Escherichia coli* ST354 isolate also contained the plasmid (**Fig 2, S4**). While short-read sequencing data alone is insufficient to provide structural data associating NDM with the plasmid backbone, the high degree of correlation between NDM and the IncF-associated plasmid genes in concert with the phylogenetic relationships between isolates strongly suggests that these genes are co-inherited (**Fig S3**). Interestingly, 9 of 17 (53%) isolates with an IncF plasmid in survey 13 were KPC+/NDM- (ST14, 1 ST258, and 1 novel), but this plasmid then became present in predominantly bla_NDM-1_ isolates, with only 0-3 isolates with the IncF plasmid being KPC+/NDM- in the subsequent surveys (0-7%) (**Fig S4**). In addition to NDM, the IncF plasmid harbored a number of antibiotic resistance genes from several different resistance classes and the qacE gene, which has been shown to provide resistance to common biocides such as chlorhexidine gluconate in *K. pneumoniae* [64] (**Fig S3**).

**Figure 2:**
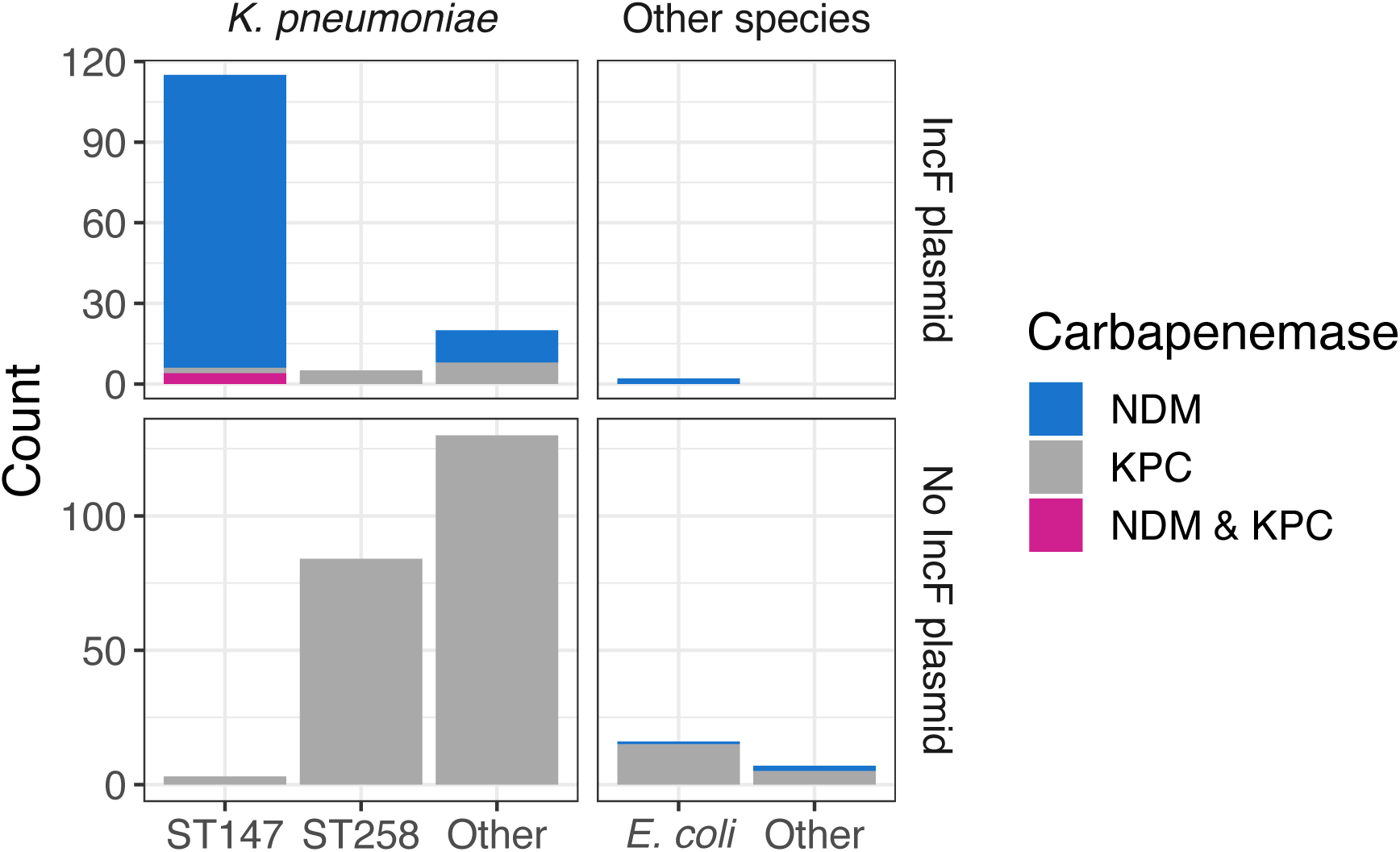
The majority of NDM+ isolates are Kp ST147 and carry NDM on an IncF plasmid. Number of sequenced isolates of various species and sequence types, what carbapenemase(s) they contain, and whether they have the IncF plasmid.

### Regional ST147 isolates are phylogenetically distinct from all public isolates

We investigated the phylogeography of ST147 in the Chicago area to determine whether circulating ST147 could be attributed to one or multiple importation events into the region. To this end we constructed a whole-genome phylogeny that included publicly available ST147 genomes from across the globe. Examination of the phylogenetic reconstruction revealed that all the ST147 isolates from the study form a monophyletic cluster, consistent with a single regional introduction (**Fig 3**). We also noted that while none of the NDM+ ST147 public isolates outside of the Chicago region harbor the IncF plasmid, the majority of ST147s from Chicago contain the plasmid. Moreover, one of the ST147 isolates in the KPC+/NDM- outgroup (from survey 13-2) contains the IncF plasmid but lacks NDM (**Fig S5**), suggesting that the plasmid may have been acquired in a locally circulating ST147 strain, followed by integration of a mobile element harboring NDM. A dated phylogenetic analysis of circulating ST147 yielded an estimate of August 2015 for when the NDM+ clade of ST147 first arose in the region (95% credible interval [CI]: February 2015 - March 2016; **Fig 4, S6; Table S2**), compared to an estimate of July 2013 for when ST147 first entered the region (CI: October 2011 – October 2014; **Table S2**).

**Figure 3:**
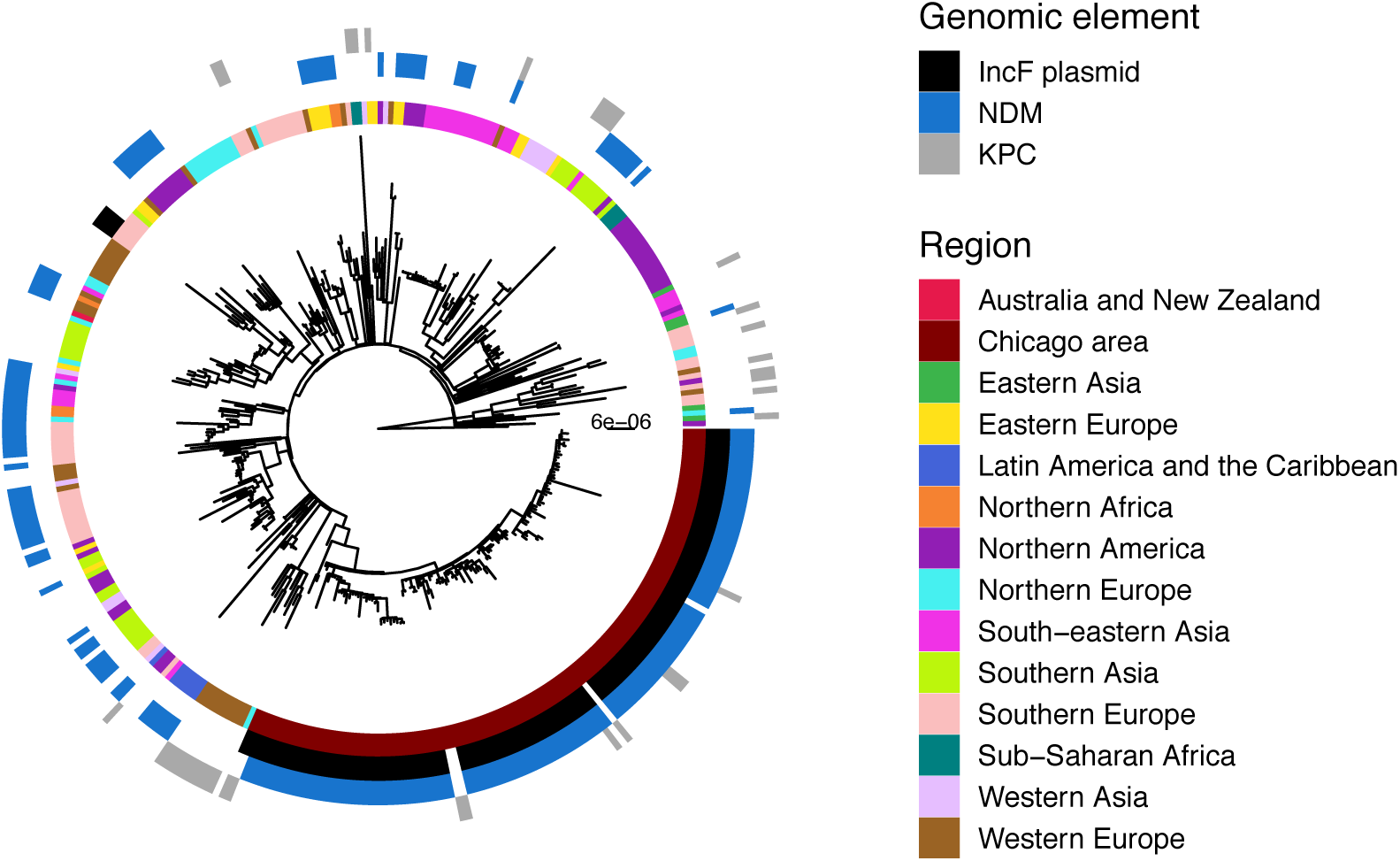
Study isolates are clonally separated from all publicly available isolates outside the Chicago region. Maximum likelihood phylogeny of study and public isolates annotated by geographic region and genomic element.

**Figure 4:**
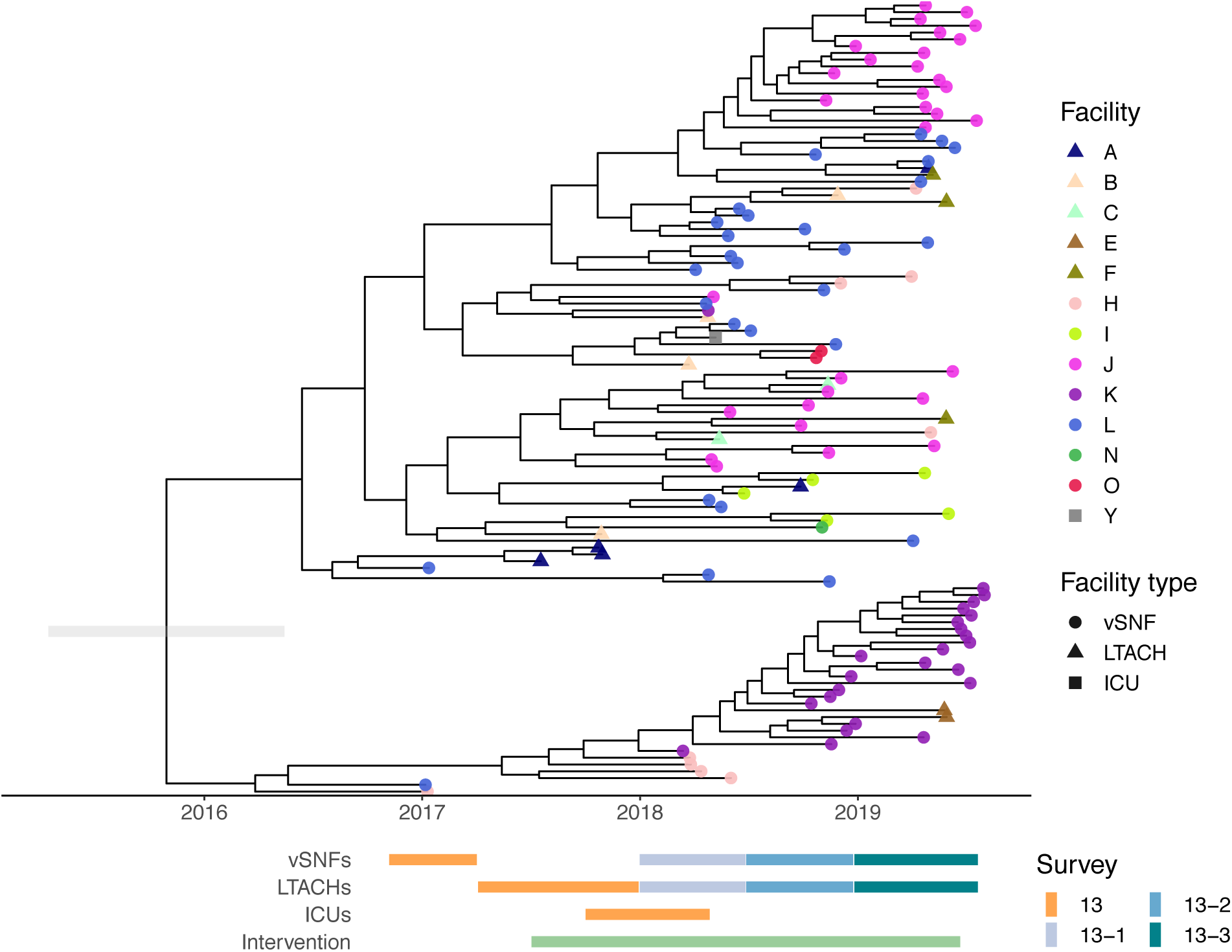
NDM+ ST147 *Klebsiella pneumoniae* was introduced into the region around 2015. Dated phylogeny generated by bactdating. Grey bar on the root is the lower and upper bounds of the confidence interval. vSNF = ventilator-capable skilled nursing facility. LTACH = long-term acute care hospital, ICU = intensive care unit.

### Genomic evidence indicates that intra-facility transmission is driving prevalence at high- prevalence vSNFs

After determining that the increase in NDM prevalence corresponds to a clonal outbreak of bla_NDM-1_ ST147, we investigated the potential transmission dynamics of this clone within and between healthcare facilities. We observed a substantial clustering of isolates from certain vSNFs on the phylogeny (**Fig 5A, S7**), which suggests potential intra-facility transmission. To further investigate whether these clusters may represent within-facility transmission, we calculated pairwise SNV distances among all pairs of isolates and compared these distances for isolates from the same facility (intra-facility pairs) to isolates from different facilities (inter-facility pairs) across surveys (**Fig 5B**). Indeed, starting in survey 13-2 we observed a disproportionate representation of small SNV distances (≤12 SNVs; see methods for threshold selection) consistent with intra-facility transmission in vSNFs. Of note, in survey 13-3 we observed spikes in small SNV distances for both intra- and inter-facility pairs, with closely related intra-facility pairs being primarily from vSNFs and closely related inter-facility pairs being from both vSNF- LTACH and vSNF-vSNF pairs (**Fig S8**). Putting these closely related inter-facility pairs in the context of the regional patient transfer network supports a potential role of patient transfer in regional bla_NDM-1_ ST147 spread in survey 13-3, with vSNF-LTACH and vSNF-vSNF isolate pairs less than 12 SNVs apart being from facilities with higher patient flow than isolate pairs with 12 or more SNVs (Wilcox p = 7.7e-6 and 1.4e-9, respectively; **Fig S9**).

**Figure 5:**
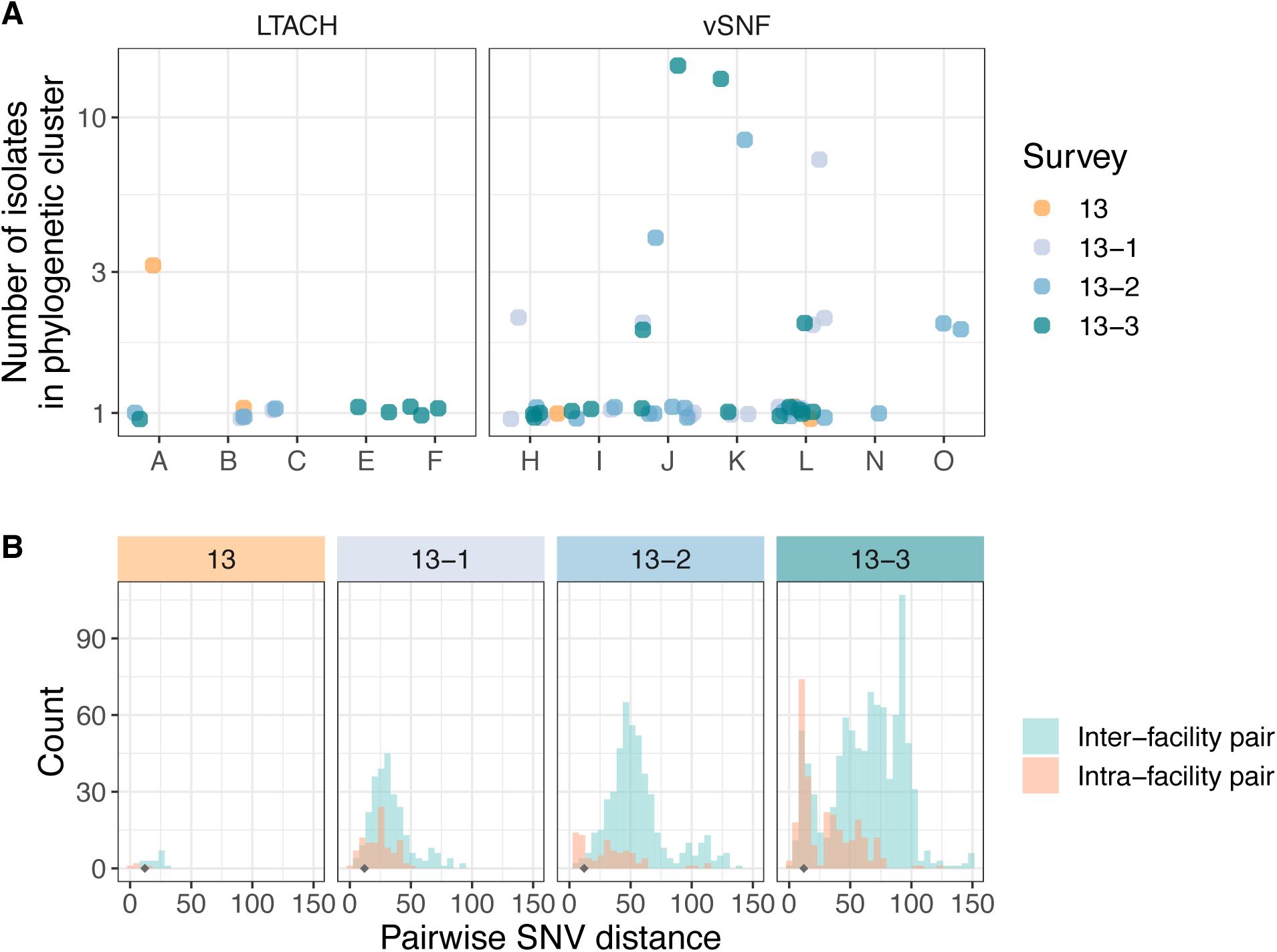
Intra-facility transmission is driving prevalence at high-prevalence vSNFs. (A) Number of isolates in the largest subclade of the maximum likelihood phylogeny containing ≥90% of isolates from the given facility (see methods for more details). (B) Pairwise SNV distances of isolates from the same and different facilities across surveys. The grey diamond at a pairwise SNV distance of 12 indicates the threshold for closely related isolates (see methods for details). vSNF = ventilator-capable skilled nursing facility. LTACH = long-term acute care hospital, ICU = intensive care unit.

## Discussion

We performed genomic analyses of CRE isolates collected through serial point-prevalence surveys in the Chicago area to understand the molecular and epidemiologic basis for an increase in NDM+ CRE prevalence across a regional healthcare network. Our analysis supports the increase in NDM+ CRE being due to the clonal dissemination of a single bla_NDM-1_ ST147 strain of *K. pneumoniae* that emerged in 2015. Putting genomic analysis in the context of the regional healthcare network supports this strain first reaching high prevalence in a small number of vSNFs due to intra-facility transmission, with potential subsequent spread to other connected healthcare facilities via patient transfer.

Whole-genome sequencing showed that the majority of bla_NDM-1_ ST147 harbored an IncF multidrug resistance plasmid. Incorporating public data into the analysis revealed that these isolates formed a monophyletic clade, suggesting a single introduction into the region, either through importation of a pre-existing NDM+ ST147 strain or acquisition of bla_NDM_ by a locally circulating ST147 strain. Furthermore, examination of the global phylogeny indicates that, while NDM+ ST147 has evolved multiple times in different locations and sometimes resulted in clonal outbreaks, none of the global NDM+ ST147 isolates we included in our analysis harbor the IncF plasmid found in our study isolates. The rapid and widespread dissemination of this strain in the region indicates that the NDM-carrying IncF plasmid we identified here can stably associate with an ST147 strain with epidemic potential. Given the potential negative impact of epidemic NDM- carrying *K. pneumoniae*, this possibility warrants close monitoring.

By combining regional surveillance with genomic analysis, we were able to discern that NDM initially spread in three vSNFs, likely via intra-facility transmission, with evidence of subsequent spread to connected healthcare facilities. There are several factors that likely contributed to the spread of this NDM+ ST147 clone. First, vSNF patients are a high-risk population for carriage of CRE as they are chronically ill, are usually admitted from ICUs or LTACHs, and are often exposed to antibiotics [65]. Second, patients in vSNFs generally have long lengths of stay — often much longer than patient stays at LTACHs [66] — meaning that they have a longer period of time to acquire a multi-drug resistant organism. Furthermore, multibed rooms are common and the facilities themselves are often under-resourced from a staffing and infection control perspective [6], both of which could facilitate intra-facility spread. Our findings paired with these observations indicate that vSNFs may be important healthcare facilities to detect emerging threats and potentially contain them before widespread dissemination. In the current study, we note that NDM+ isolates were uncommon in ICUs, and the outbreak of ST147 might not have been detectable until much later if sampling were restricted to ICUs.

Of note, the Chicago PROTECT intervention, a bundled infection prevention intervention designed to reduce the prevalence of CRE, began at around the same time as NDM+ ST147 began amplifying in the region. There are several potential reasons for this association. The intervention could have created a new niche for NDM+ isolates to thrive in, either due to NDM itself, or other genes on the associated plasmid. One possibility is that the qacE gene harbored on the NDM IncF plasmid might have provided a selective advantage in the context of an intervention that included chlorhexidine gluconate bathing. It is also possible that the emergence of NDM during the intervention was coincidental, and that the intervention might have even prevented a larger NDM+ outbreak from occurring across more facilities. Regardless, this study exemplifies how genomic surveillance can be used to monitor emerging threats and understand how they develop and circulate through a region.

Our study has several strengths. Active surveillance of diverse types of healthcare facilities in the region allowed us to identify and investigate a potential multi-drug resistant organism threat earlier than would have been possible if serial point-prevalence surveys across several facility types were not ongoing. Furthermore, cross-sectional patient sampling within each survey allowed us to obtain a complete snapshot of CRE prevalence at a given facility at a given point in time, and to detect the increase in NDM+ isolates over time. Finally, we were able to leverage information from whole-genome sequencing to investigate the relatedness of isolates, as well as the intra- and inter-facility transmission dynamics of NDM across the healthcare network.

Our study also has several important limitations. First, we only have data from discrete points in time for each survey. This could have led to potential biases in the number of NDM+ isolates sequenced at facilities given that the patients at these facilities had different average lengths of stay, and it also precluded more nuanced examination of intra-facility transmission dynamics. Second, we lack data from short-term acute care or community settings, particularly in the last three surveys, which limited our ability to examine the relative importance of other regional reservoirs for NDM+ ST147. However, the short-term acute care data that is available did not support their role in bla_NDM-1_ ST147 expansion. Another limitation is that we only had access to short-read sequencing data, which limited our ability to investigate more complex plasmid dynamics. While we plan to perform long-read sequencing on a subset of these isolates in the future, we find it notable that we were able to leverage publicly available complete plasmid sequences to determine that NDM was carried on the same plasmid in the majority of isolates.

In conclusion, we were able to combine whole-genome sequencing of NDM+ isolates from serial point-prevalence surveys at vSNFs, LTACHs, and ICUs to identify and investigate the dissemination of an NDM+ clone of *K. pneumoniae*. These findings highlight the importance of performing regional point-prevalence surveys in not only acute care hospital ICUs, but also in post-acute care LTACHs and vSNFs, to identify potential multi-drug resistant organism threats as early as possible. Furthermore, our study identified an emerging bla_NDM-1_ ST147 clone of *K. pneumoniae* that could pose a threat to healthcare facilities worldwide if this plasmid/strain combination continues to spread.

## Data Availability

All sequenced isolates will be deposited under BioProject PRJNA686897.
Code for analysis and visualization, as well as supplementary data, can be found here: https://github.com/Snitkin-Lab-Umich/ndm-st147-chicago-ms.

https://www.ncbi.nlm.nih.gov/bioproject/?term=prjna686897

https://github.com/Snitkin-Lab-Umich/ndm-st147-chicago-ms

## Acknowledgements

We gratefully acknowledge the patients and staff of the facilities for their participation in this study. We thank officials from Chicago Department of Public Health, Cook County Department of Public Health, Illinois Department of Public Health, and Centers for Disease Control and Prevention for their direct involvement in and support of Chicago PROTECT. We also thank Louis Fogg and Vincent Young for fruitful discussion of the analyses presented here. We thank Ellen Benson, Mary Carl Froilan, Claire Heshmat, Jinal Makhija, and Mitali Shah for their role in specimen and data collection at participating healthcare facilities, and Pamela Bell and Karen Lolans for their role in laboratory analysis.

## Authors’ contributions

All authors developed methodology and reviewed and edited the manuscript. ML, ES, MH, AMJ, and ZL conceptualized the research goals and aims. ZL, RC, AP, and ES developed and implemented software and curated the data. ZL, RC, and AP performed formal analysis. ML, MH and ES provided resources. ZL and ES wrote the original draft. ZL and RC visualized the results. ML, MH, AMJ, and ES provided supervision. ML, MH, and ES managed the project. ML, MH, and ES acquired funding.

## Supplementary material

### Supplementary methods

#### Public isolates

All Klebsiella genomes available as of October 15, 2020 in the PATRIC database [30] were downloaded. Furthermore, we included additional whole-genome sequences of NDM+ ST147 genomes identified through a literature search [31–39]. Information about the public genomes included can be found in supplementary data file 2. Complete genomes and any ST147 genome were used for downstream analysis. For isolates where only genome assemblies were available, we used wgsim v1.9 [40] to simulate reads of length 250 from the assemblies with no base error rate, rate of mutation and fraction of indels. These simulated reads were used to perform variant calling and generate a phylogenetic tree (see below).

#### Multi-locus sequence typing

Multi-locus sequence types (MLSTs) were determined with MLSTyper [13] for public isolates and with ARIBA v2.14.5 for study isolates [14].

#### Assembly generation and annotation

For all study isolates, the reads were assembled using assemblage v1.2 [15], which runs the following steps sequentially: down-sample reads to 150X coverage using Seqtk v1.3 [16] and Mash v2.2.2 [17], clean down-sampled reads with Trimmomatic v0.39 [12], assemble clean reads with SPAdes v3.14.1 [18], perform post-assembly correction with PILON v1.23 [19], and remove contigs smaller than 500bp. Genomes were annotated with RAST v1.3.0 [20]. The quality of genomes was then assessed by the number of coding sequences and any genomes containing fewer than 4000 or greater than 8000 were excluded from the analysis.

#### Sequencing quality control and ST147 single nucleotide variant identification

The quality of sequencing reads was assessed with FastQC v0.11.9 [11], and Trimmomatic v0.39 [12] was used for trimming adapter sequences and low-quality bases. Single nucleotide variants (SNVs) were identified by (i) mapping filtered reads to the ST147 Kp46596 reference genome (Genbank accession NZ_CP059312.1) using the Burrows-Wheeler short-read aligner (bwa v0.7.17 [21]), (ii) discarding polymerase chain reaction duplicates with Picard v2.24.1 [22], and (iii) calling variants with SAMtools and bcftools v1.9 [23]. Variants were filtered from raw results using VariantFiltration from GATK v4.1.9.0 [24] (QUAL >100; MQ >50; >=10 reads supporting variant; and FQ < 0.025). In addition, a custom python script was used to filter out (mask) single nucleotide variants in the whole-genome alignment that were: (i) <5 base pairs (bp) in proximity to indels that were identified by GATK HaplotypeCaller [25], (ii) in a recombinant region identified by Gubbins v2.3.4 [26], in a phage region identified by the Phaster web tool [27] or (iii) they resided in tandem repeats of length greater than 20bp as determined using the exact-tandem program in MUMmer v3.23 [28].

Genomes were removed that showed evidence of contamination as inferred by poor mapping to the reference genome or excessive numbers of uniquely filtered variants. This whole-genome masked variant alignment was used to reconstruct a maximum likelihood phylogeny with IQ-TREE v1.6.12 [41] using the general time reversible model GTR+G and ultrafast bootstrap with 1000 replicates (-bb 1000) [42].

#### Identifying NDM-containing plasmids

To identify the presence of beta-lactamase containing plasmids, we searched for previously sequenced, closed plasmids which were known to harbor genes of interest. First, CD-HIT v4.7 was used to identify orthologous genes [47]. NDM or KPC containing contigs from complete genomes were identified by the presence of a gene annotated as either of these beta-lactamases in the gff file, and all genes contained on these contigs were identified. The plasmid sharing the maximum number of genes with the NDM+ isolates was identified. Presence of the plasmid was determined as sharing 80% of the shared plasmid genes. A core gene alignment was created with cognac [46]. The pairwise substitution distance matrix was calculated from the alignment, which was then used to create a neighbor joining tree using ape v5.3 [29] and midpoint rooted using phytools v0.6-99 [48]. The rows of the gene presence absence matrix were then sorted by the tip labels on the neighbor joining tree.

To confirm the presence of NDM containing plasmid, which was prevalent in the ST147 isolates, we utilized the raw sequencing data to calculate coverage across the plasmid sequence. We selected the complete genome with the best match for the NDM+ plasmid by shared gene content across the plasmid sequence as the reference sequence. The sequencing reads were aligned to the plasmid sequence using bwa mem v0.7.15 [21]. The sam files were parsed to identify reads which aligned to the NDM plasmid, and coverage was then calculated for 500 bp bins across the plasmid sequence. Antibiotic resistance genes were identified by querying the CARD database [49] with BLAST [50].

#### Inferring ancestral dates

We used the R package BactDating v1.0.12 [43] to infer ancestral dates on the NDM+ clade of the ST147 maximum likelihood phylogeny, as well as the phylogeny including all study ST147 isolates. A root-to-tip analysis was first performed and indicated that there is temporal signal in the data. As we do not have access to actual isolate dates, a date range for each tree tip was determined based on the year of isolation as well as the survey and facility type that the isolate came from. The maximum likelihood phylogeny and the date ranges were used to infer ancestral dates. All available molecular clock models were tested, each run for 1,000,000 MCMC iterations. The R package coda v0.19-4 [45] was used to evaluate MCMC convergence. All models had similar estimated root dates for the NDM+ clade (range: 2013.96 to 2016.24), and slightly more variation for the ST147 isolates (1996 to 2014), with varying levels of uncertainty. The best fit models were the additive relaxed clock (ARC) family of models [44], with the best fit being mixed continuous ARC (cARC) for the NDM+ clade and ARC for all ST147 study isolates. The cARC model was chosen for all subsequent analyses with the NDM+ clade, and the ARC model was chosen for all subsequent analyses with the ST147 study isolates. The effective sample size for all parameters in the best models were above 400.

#### Identifying phylogenetic clustering

We subset the NDM+ clade of the ST147 maximum likelihood phylogeny by survey. For each survey-specific tree, we used a method modified from [8] to identify the largest subclades that contain ≥90% of isolates from the same facility.

#### Calculating pairwise SNV distances

Pairwise SNV distances were calculated for the gubbins recombination-filtered ST147 variant alignment using the dist.dna function in the R package ape v5.4-1 [29], with the “N” model (count), and pairwise deletion set to true to include both core and non-core genetic variation. We chose a pairwise SNV distance threshold based on the BactDating estimated mutation rate of ∼12 SNVs per year (**Table S2**). As the surveys were conducted 6 months apart, we considered closely related isolate pairs to be those with a SNV distance of ≤12 (i.e. 6 mutations per isolate). Wilcox tests were used to calculate significance, and all p-values were Bonferroni- corrected.

#### Determining paths of maximum patient flow

The patient transfer network consists of healthcare facility nodes and patient transfer edges. A patient transfer includes direct transfers of no more than one day between discharge from source facility and admission to destination facility, and indirect transfers of patients with an intervening stay in the community or an intermediate facility between discharge and readmission. Normalized edge weights were calculated by dividing the number of patient transfers from source facility to destination facility in 2016 (i.e. within 365 days) by the total number of outgoing patient transfers at the source facility. We calculated patient flow as the product of the normalized edge weights for the path of maximum patient flow [8]. R igraph v1.2.6 was used for all patient flow analyses [51].

## Supplementary Figures

**Figure S1:**
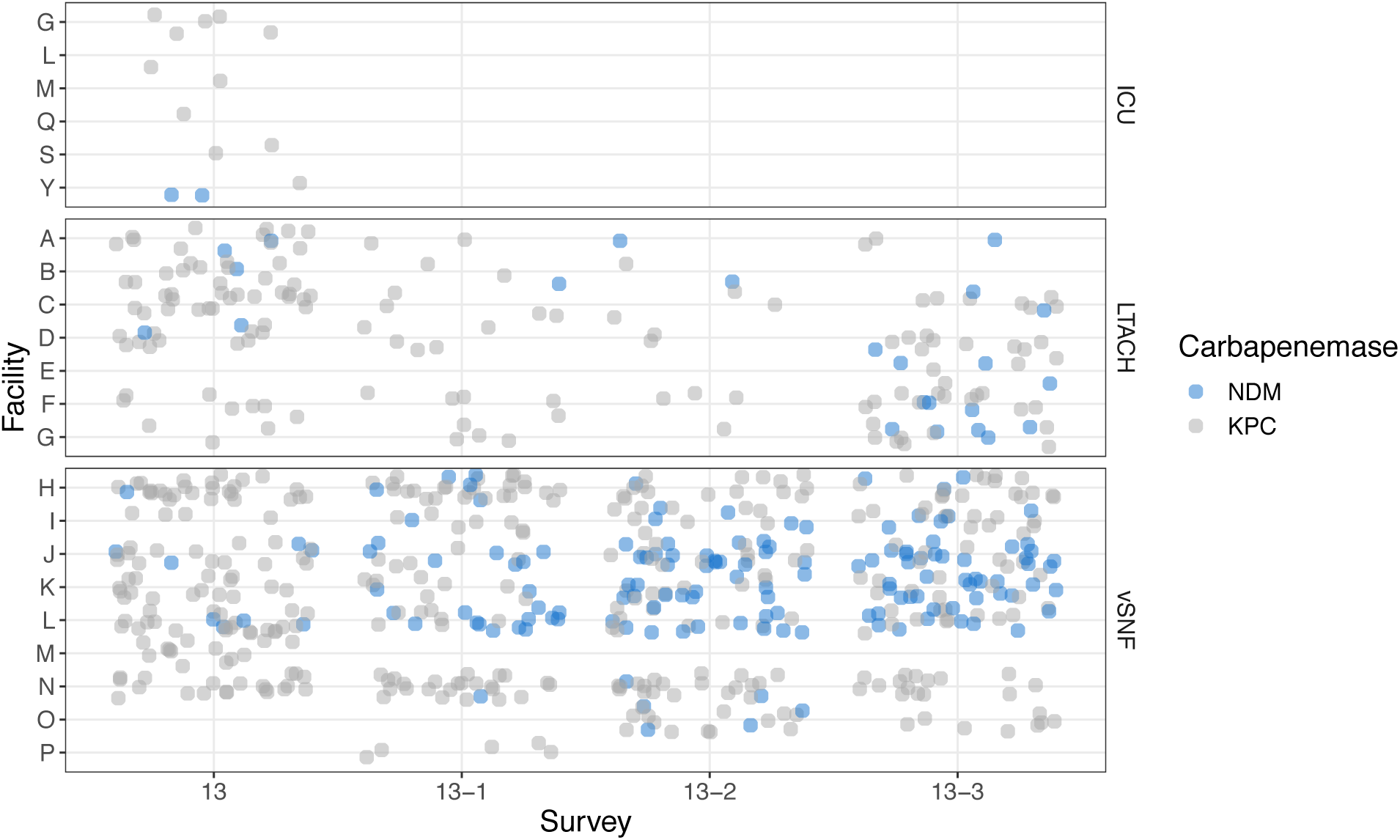
NDM counts increase over time in some vSNFs and LTACHs. vSNF = ventilator-capable skilled nursing facility. LTACH = long-term acute care hospital, ICU = intensive care unit.

**Figure S2:**
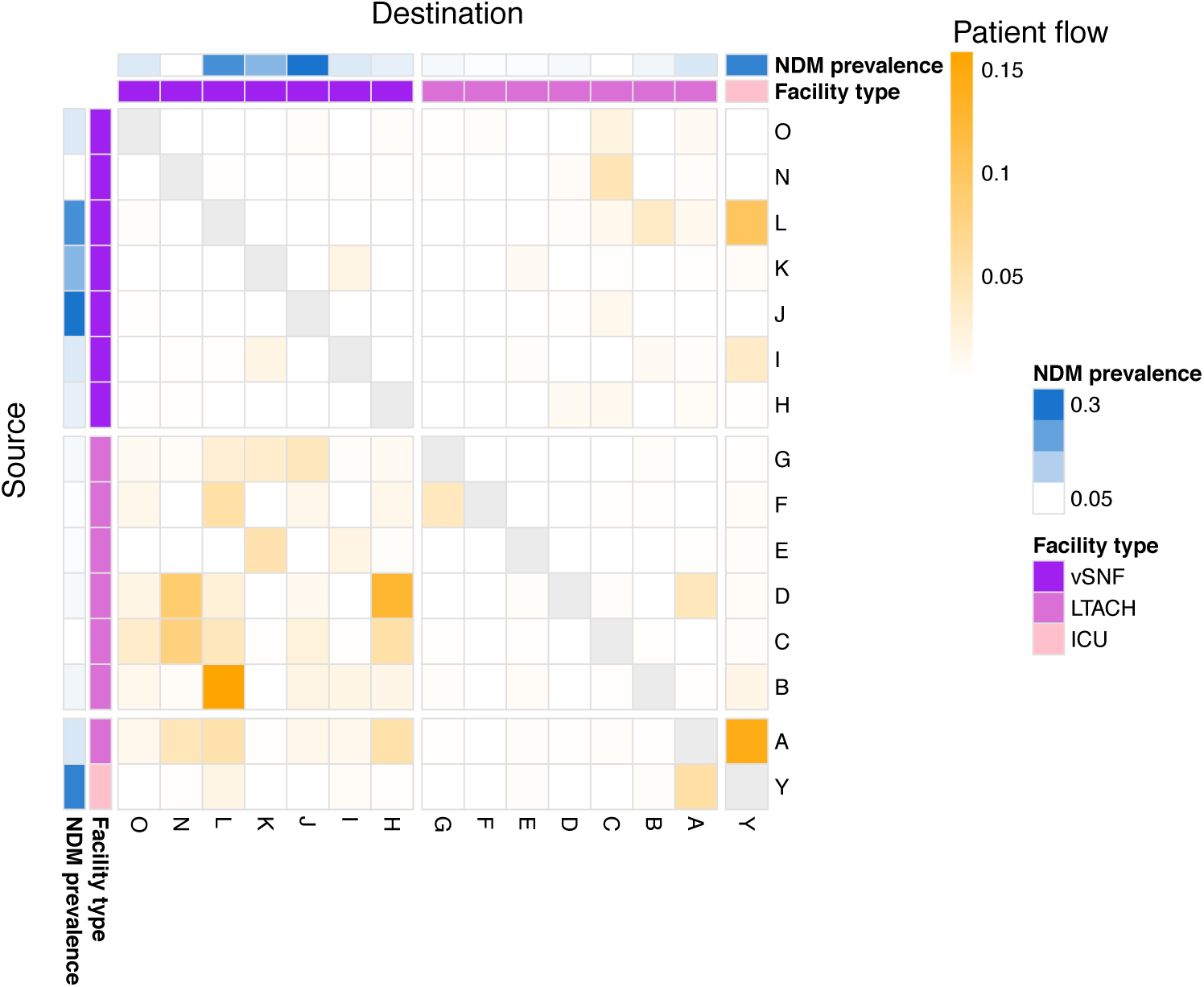
High patient flow into and/or out of a facility does not correlate with high NDM prevalence. See methods for patient flow calculation. vSNF = ventilator-capable skilled nursing facility. LTACH = long-term acute care hospital, ICU = intensive care unit.

**Figure S3:**
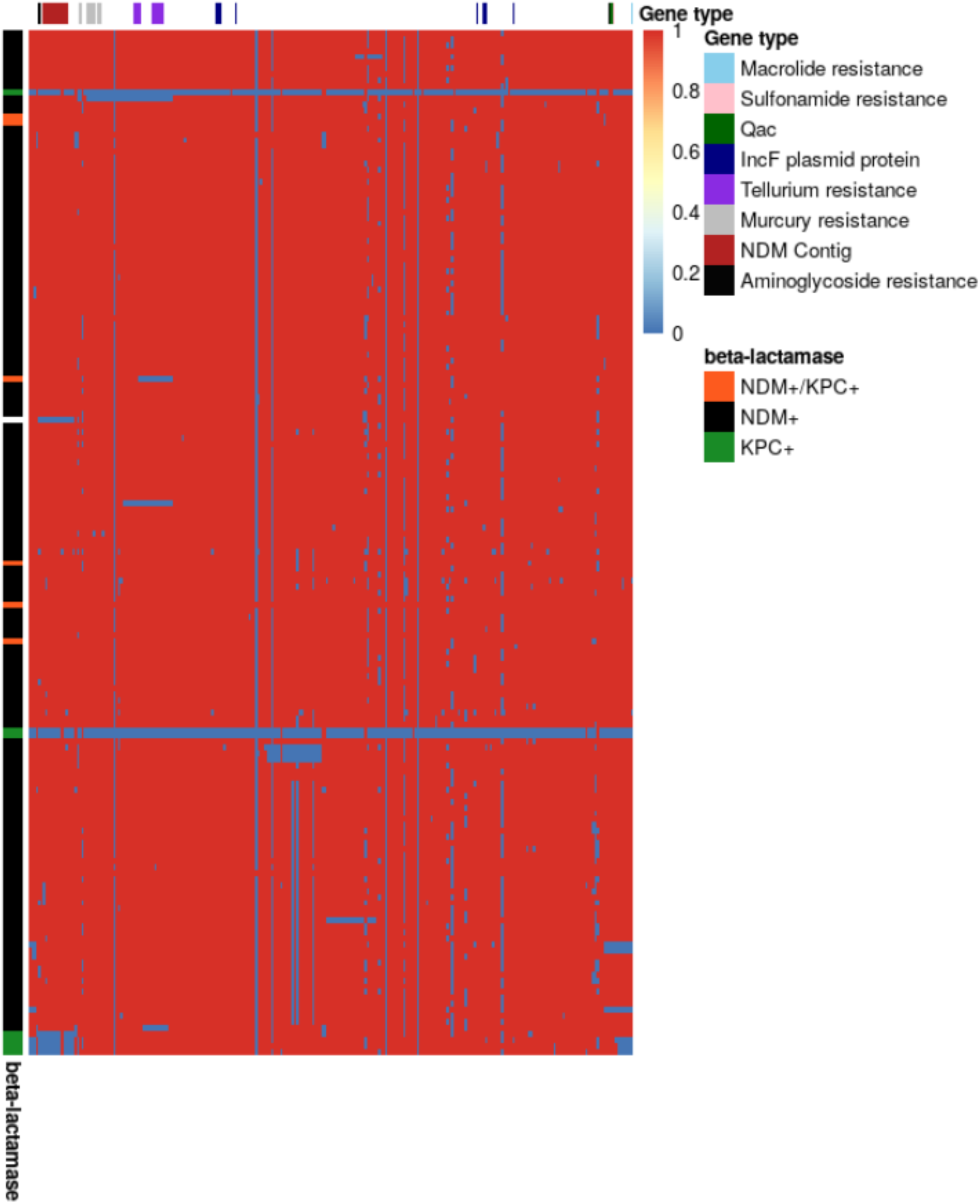
The majority of *K. pneumoniae* ST147 isolates from this study contain the majority of genes from a multi-drug resistant IncF plasmid, including an NDM-/KPC+ outgroup. Heatmap of genes on plasmid rows are genome sequences and columns are genes. Rows are ordered by the phylogeny and columns are linear order of the genes on the complete plasmid sequence. Red indicates gene presence and blue indicates gene absence. Loci of interest are indicated above the heatmap.

**Figure S4:**
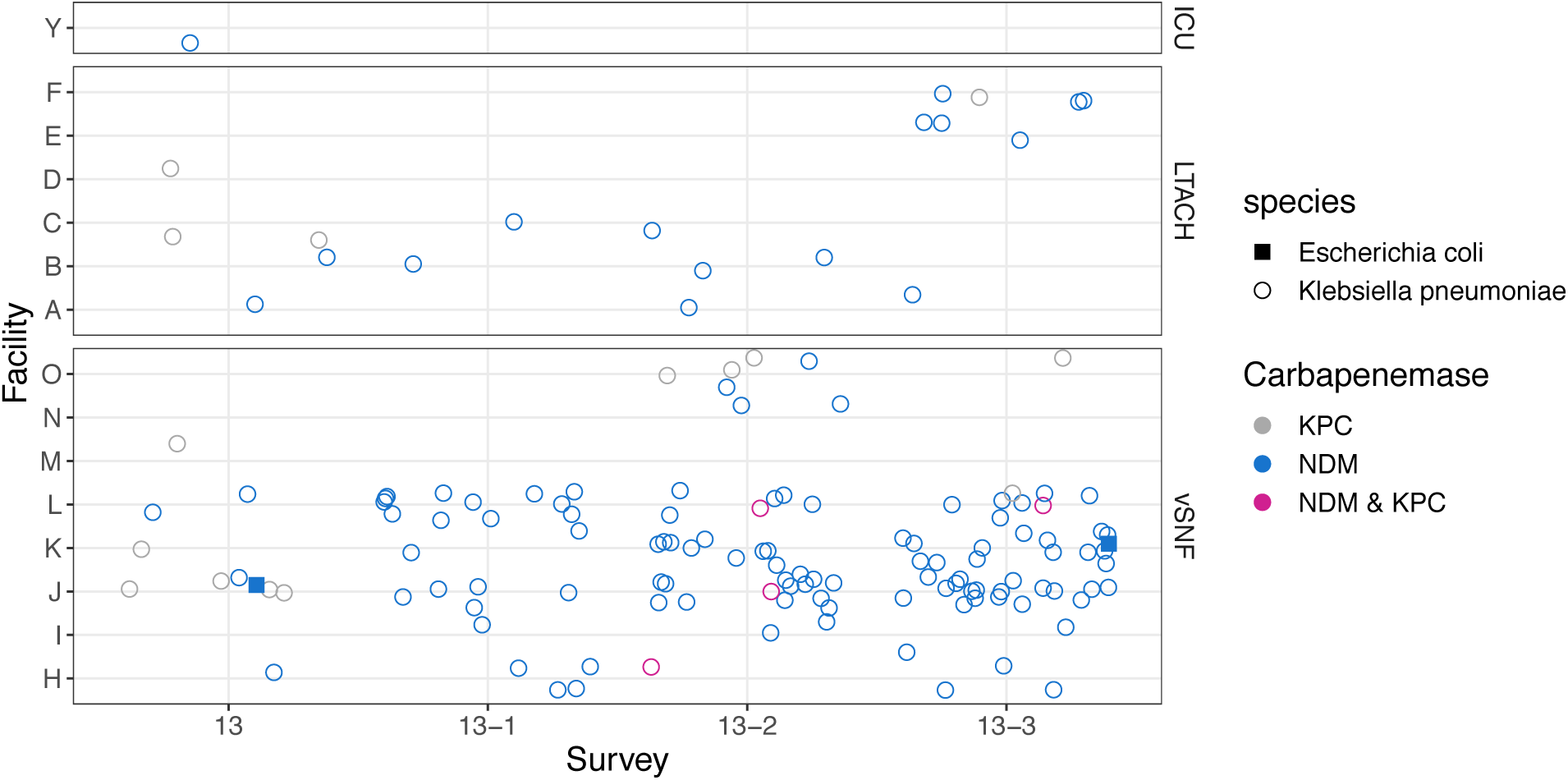
The IncF plasmid is present in KPC+ isolates early on, but then becomes predominantly present in NDM+ isolates. Only isolates with the IncF plasmid are depicted here. vSNF = ventilator-capable skilled nursing facility. LTACH = long-term acute care hospital, ICU = intensive care unit.

**Figure S5:**
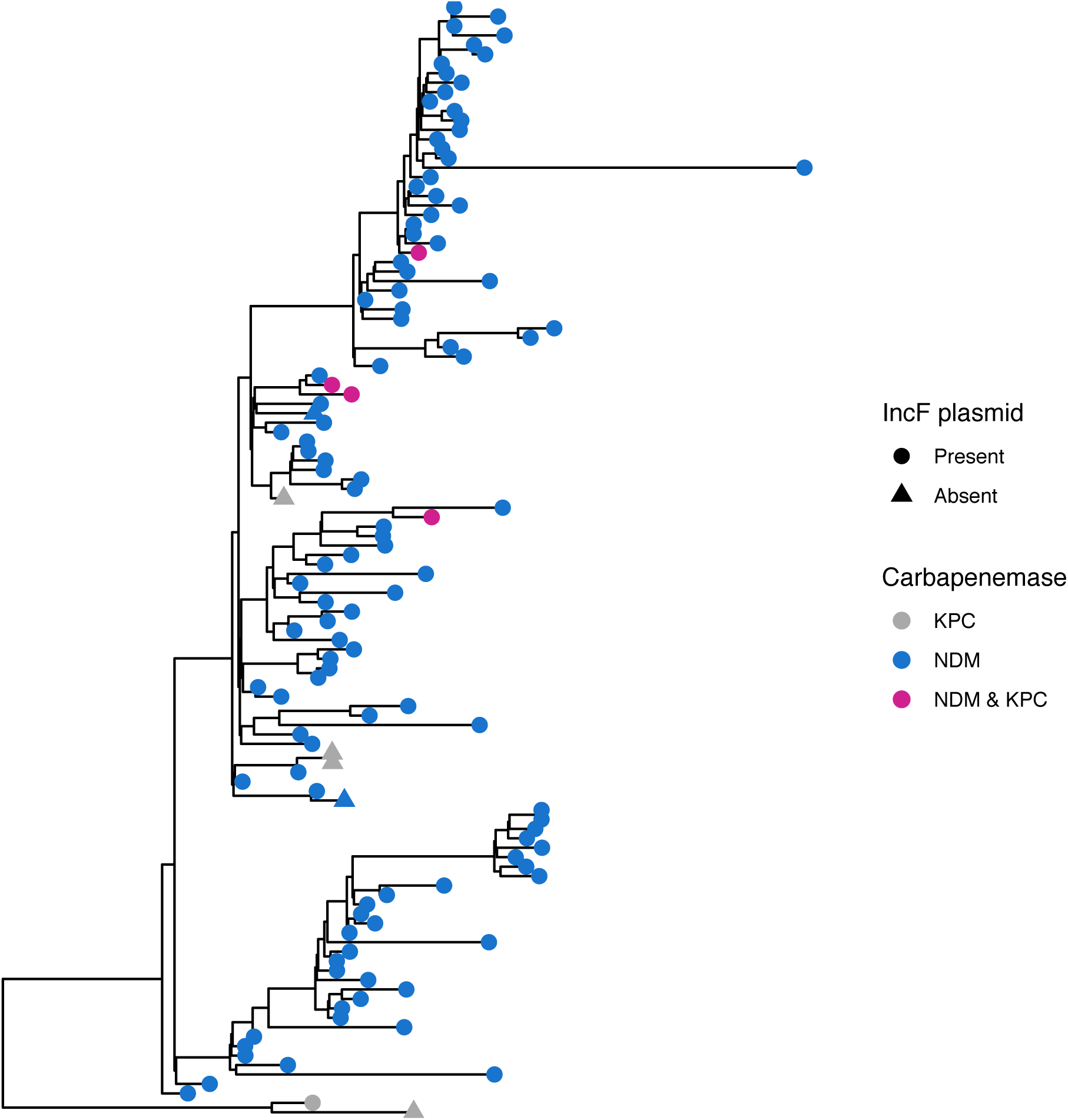
the IncF plasmid is present in a KPC+ outgroup of the NDM+ ST147 clade. Maximum likelihood phylogeny of study ST147 isolates annotated by IncF plasmid presence, KPC, and NDM.

**Figure S6:**
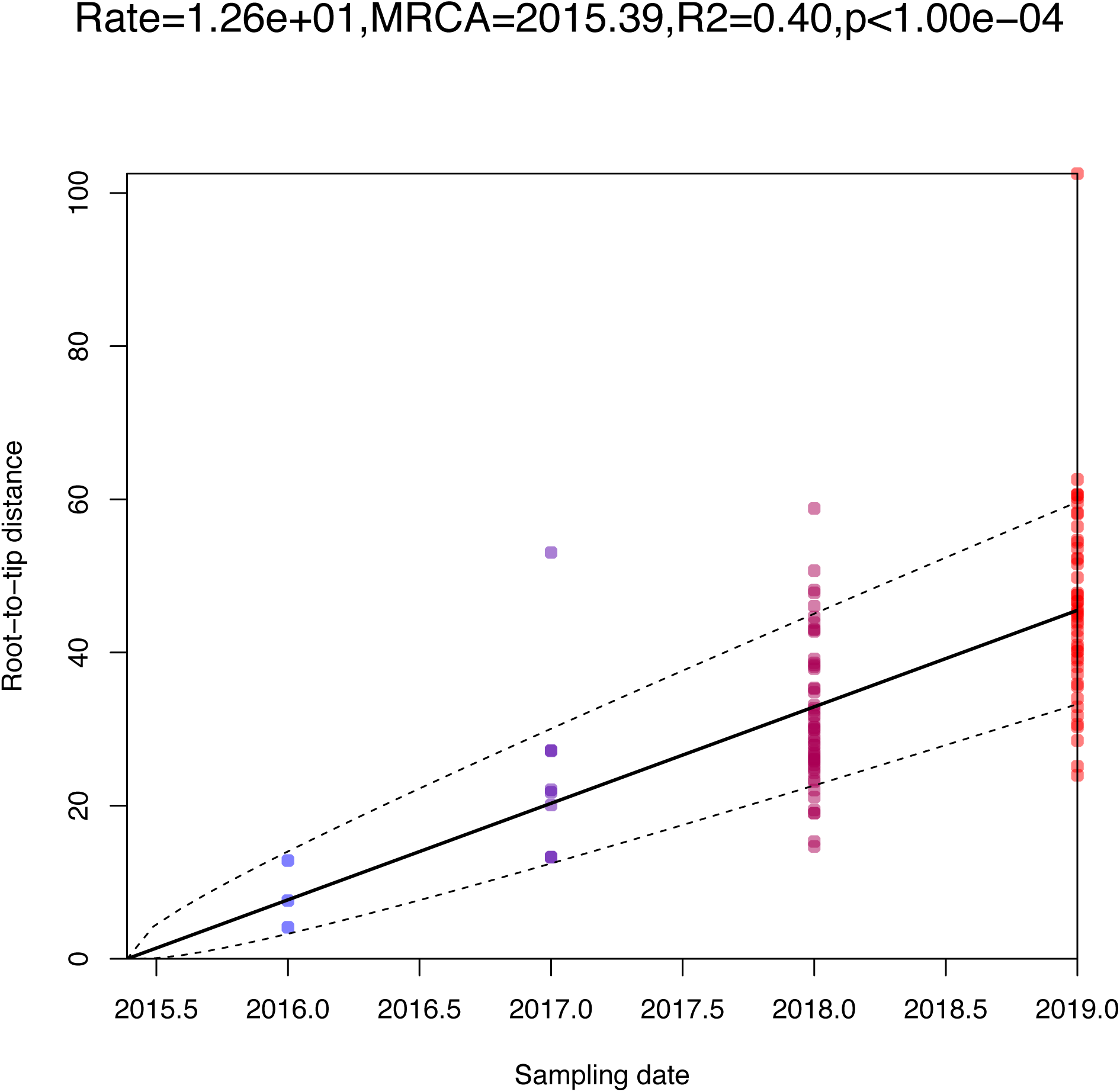
Root-to-tip distance of study isolates reveals strong temporal signal.

**Figure S7:**
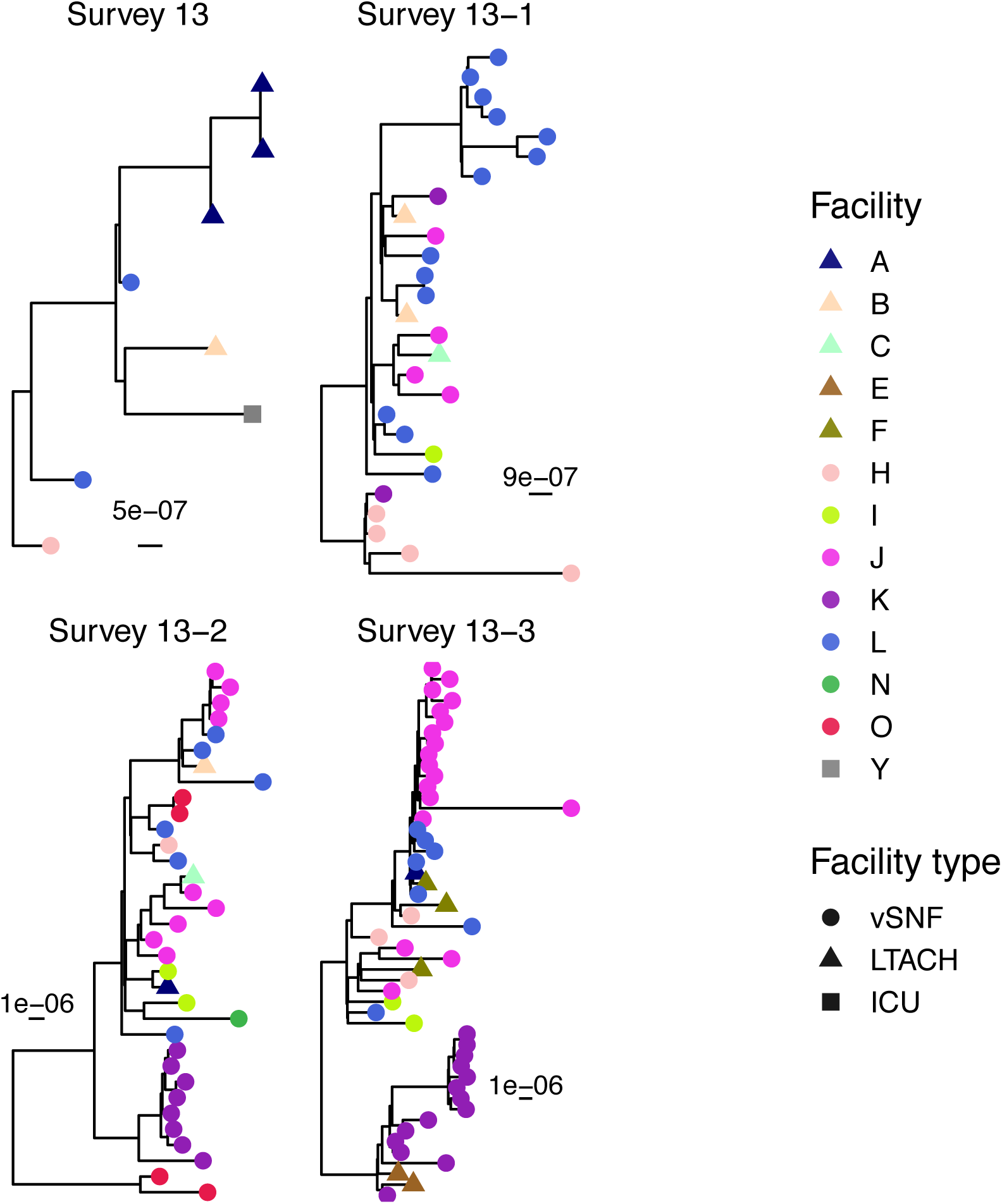
Isolates from a given vSNF often cluster on the phylogeny. Maximum likelihood phylogenies labeled by facility for each survey. These phylogenies were used to identify the phylogenetic clusters depicted in Figure 5A. vSNF = ventilator-capable skilled nursing facility. LTACH = long-term acute care hospital, ICU = intensive care unit.

**Figure S8:**
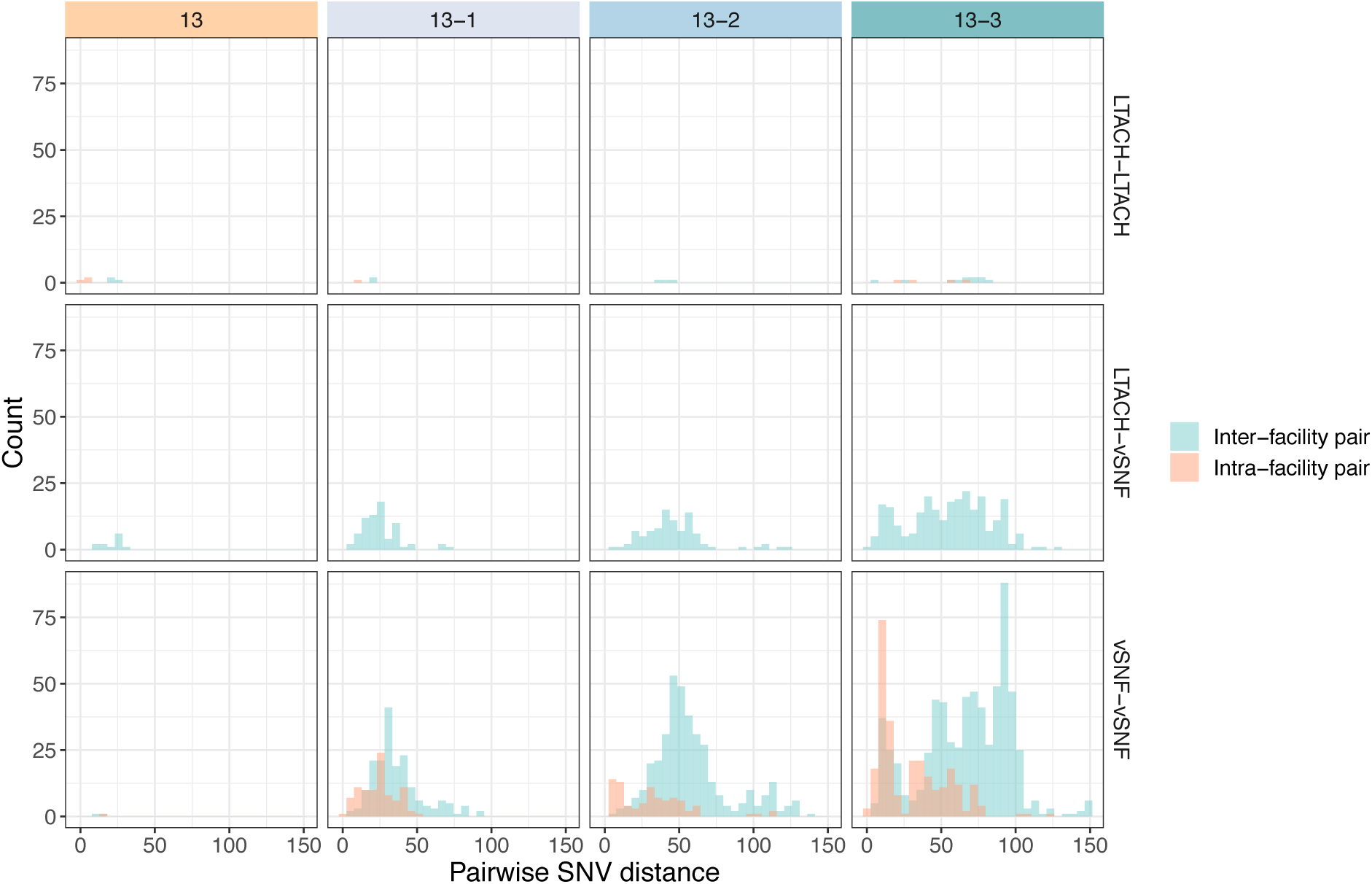
The majority of closely related intra-facility isolate pairs are from vSNFs, while there are closely related inter-facility vSNF-vSNF pairs and LTACH-vSNF pairs. vSNF = ventilator-capable skilled nursing facility. LTACH = long-term acute care hospital.

**Figure S9:**
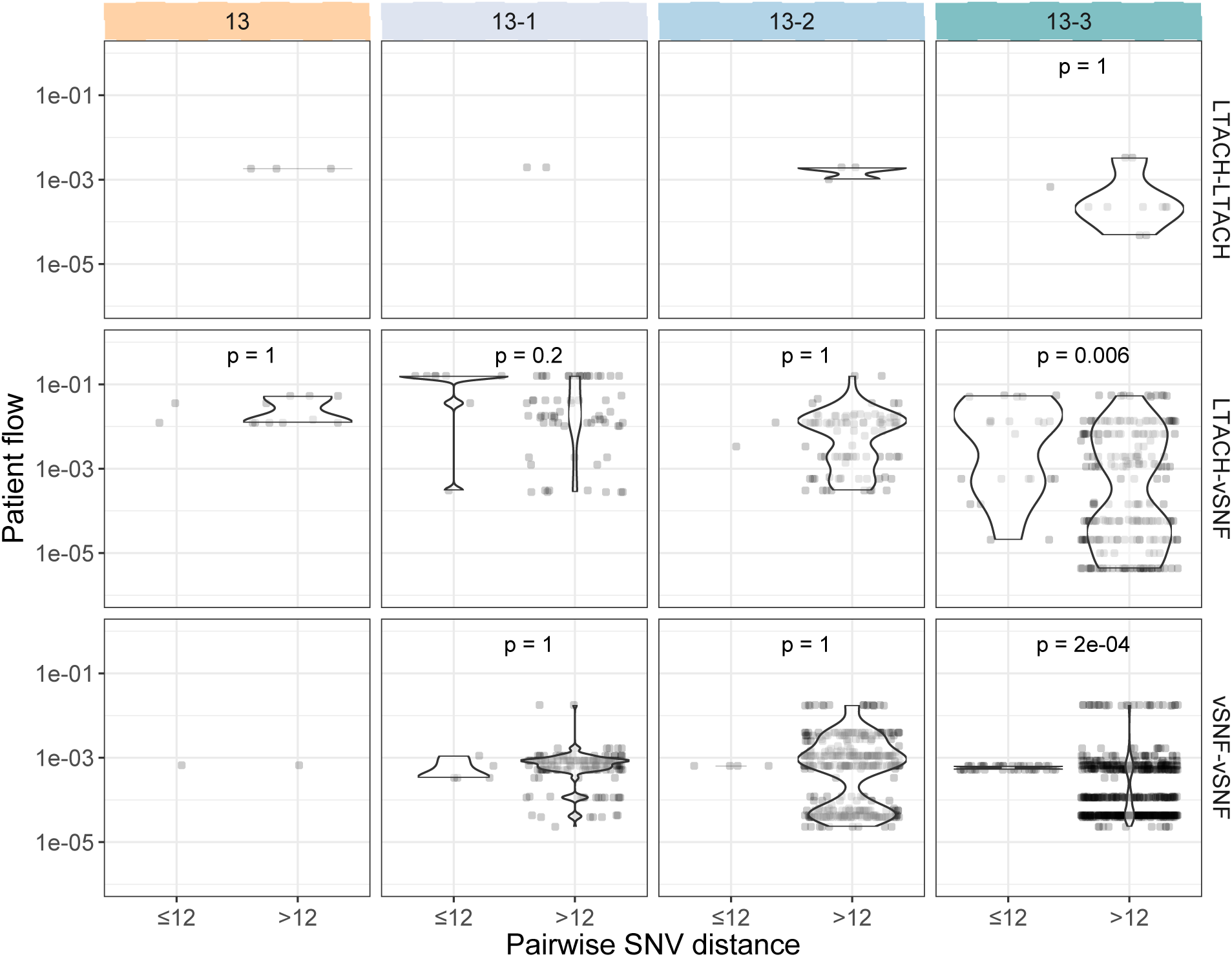
Increasing signal of transmission via the patient transfer network over time. Patient flow for inter-facility isolate pairs within 20 SNVs and not within 20 SNVs for different facility pairs over time. See methods for details on patient flow calculation. Bonferroni-adjusted p-values were calculated using the Wilcox test. vSNF = ventilator-capable skilled nursing facility. LTACH = long-term acute care hospital.

## Supplementary Tables

**Table S1:** Counts and prevalence of NDM and KPC in surveys 13 and 13-3.

| Facility | Survey 13 |  |  |  |  | Survey 13-3 |  |  |  |  | Fisher's exact p |  |
| --- | --- | --- | --- | --- | --- | --- | --- | --- | --- | --- | --- | --- |
|  | NDM |  | KPC |  | Total | NDM |  | KPC |  | Total |  |  |
|  | Count | Proportion | Count | Proportion | Count | Count | Proportion | Count | Proportion | Count | NDM | KPC |
| vSNF K | 0 | 0.00 | 16 | 0.34 | 47 | 16 | 0 | 9 | 0 | 48 | 8.2e-05 | 0.33 |
| vSNF J | 4 | 0.10 | 15 | 0.38 | 40 | 18 | 0 | 7 | 0 | 37 | 1.7e-03 | 0.33 |
| vSNF L | 4 | 0.08 | 22 | 0.45 | 49 | 12 | 0 | 14 | 1 | 35 | 1.7e-02 | 0.89 |
| vSNF I | 0 | 0.00 | 4 | 0.09 | 47 | 5 | 0 | 13 | 0 | 45 | 7.5e-02 | 0.18 |
| LTACH C | 0 | 0.00 | 19 | 0.30 | 63 | 2 | 0 | 7 | 1 | 36 | 3.1e-01 | 0.82 |
| LTACH G | 0 | 0.00 | 3 | 0.06 | 48 | 5 | 0 | 8 | 0 | 73 | 3.2e-01 | 0.89 |
| LTACH F | 0 | 0.00 | 7 | 0.15 | 48 | 3 | 0 | 11 | 0 | 66 | 4.5e-01 | 0.89 |
| vSNF H | 1 | 0.02 | 27 | 0.43 | 63 | 3 | 0 | 16 | 1 | 43 | 4.5e-01 | 0.89 |
| LTACH A | 2 | 0.06 | 11 | 0.34 | 32 | 1 | 0 | 6 | 0 | 23 | 1.0e+00 | 0.89 |
| LTACH D | 2 | 0.04 | 11 | 0.22 | 50 | 1 | 0 | 9 | 0 | 39 | 1.0e+00 | 1.00 |
| LTACH B | 1 | 0.02 | 12 | 0.24 | 49 | 0 | 0 | 2 | 0 | 31 | 1.0e+00 | 0.33 |
| vSNF N | 0 | 0.00 | 20 | 0.34 | 59 | 0 | 0 | 15 | 0 | 51 | 1.0e+00 | 0.89 |
| ICU Y | 2 | 0.29 | 1 | 0.14 | 7 | NA | NA | NA | NA | NA | NA | NA |
| vSNF O | 4 | 0.12 | 13 | 0.39 | 33 | NA | NA | NA | NA | NA | NA | NA |
| LTACH E | 0 | 0.00 | 0 | 0.00 | 55 | NA | NA | NA | NA | NA | NA | NA |
| ICU G | NA | NA | 5 | 0.38 | 13 | NA | NA | NA | NA | NA | NA | NA |
| ICU L | NA | NA | 1 | 0.12 | 8 | NA | NA | NA | NA | NA | NA | NA |
| ICU M | NA | NA | 1 | 0.10 | 10 | NA | NA | NA | NA | NA | NA | NA |
| vSNF M | NA | NA | 13 | 0.37 | 35 | NA | NA | NA | NA | NA | NA | NA |
| ICU Q | NA | NA | 1 | 0.04 | 24 | NA | NA | NA | NA | NA | NA | NA |
| ICU S | NA | NA | 2 | 0.20 | 10 | NA | NA | NA | NA | NA | NA | NA |

**Table S2:** Model convergence (effective sample sizes; ESSs), parameter estimates, and model fit (Deviance Information Criterion DIC) for dated trees.

| ST147 clade | Model | $\mu$ ESS | $\sigma$ ESS | $\alpha$ ESS | Clock rate | Root date | Root date lower CI | Root date upper CI | DIC |
| --- | --- | --- | --- | --- | --- | --- | --- | --- | --- |
| NDM+ | arc | 494.55 | 501.00 | 501.00 | 12.35 | 2015.64 | 2014.97 | 2016.21 | 1660.39 |
| NDM+ | carc | 501.00 | 501.00 | 501.00 | 12.37 | 2015.73 | 2015.15 | 2016.24 | 1664.36 |
| NDM+ | mixedcarc | 527.75 | 501.00 | 501.00 | 12.31 | 2015.72 | 2015.09 | 2016.17 | 1652.96 |
| NDM+ | mixedgamma | 415.68 | 514.12 | 425.03 | 15.85 | 2014.65 | 2008.40 | 2016.18 | 1774.02 |
| NDM+ | negbin | 392.70 | 170.49 | 437.98 | 13.73 | 2013.96 | 2006.93 | 2016.07 | 1848.19 |
| NDM+ | poisson | 298.69 | 0.00 | 405.98 | 14.99 | 2016.24 | 2015.94 | 2016.49 | 1880.35 |
| NDM+ | relaxedgamma | 434.05 | 501.00 | 603.26 | 15.70 | 2014.95 | 2012.24 | 2016.16 | 1776.49 |
| NDM+ | strictgamma | 406.80 | 0.00 | 396.89 | 11.53 | 2016.02 | 2015.67 | 2016.30 | 1726.09 |
| Entire | arc | 501.00 | 467.65 | 501.00 | 12.25 | 2013.64 | 2011.86 | 2014.91 | 1699.41 |
| Entire | carc | 409.65 | 501.00 | 501.00 | 12.28 | 2013.76 | 2012.35 | 2015.06 | 1734.89 |
| Entire | mixedcarc | 501.00 | 501.00 | 501.00 | 12.36 | 2013.82 | 2012.41 | 2015.02 | 1735.82 |
| Entire | mixedgamma | 412.99 | 434.88 | 373.22 | 15.35 | 2010.78 | 1999.35 | 2014.94 | 1746.24 |
| Entire | negbin | 434.78 | 501.00 | 305.49 | 13.57 | 2009.23 | 1996.81 | 2014.25 | 1844.46 |
| Entire | poisson | 325.67 | 0.00 | 297.94 | 15.02 | 2014.75 | 2014.03 | 2015.37 | 1981.59 |
| Entire | relaxedgamma | 412.08 | 421.97 | 422.49 | 15.29 | 2010.58 | 1996.22 | 2014.77 | 1827.66 |
| Entire | strictgamma | 383.56 | 0.00 | 399.56 | 11.74 | 2013.91 | 2013.02 | 2014.71 | 1731.43 |

## Supplementary data

1. Spreadsheet with isolate id, patient id, survey, survey year, facility, NDM/KPC, species, ST, plasmid.
2. Spreadsheet with metadata on public isolates.

